# Biallelic *IRAK4* Variants Associated with Severe Neurological Autoinflammation: An Expansion of the Clinical Phenotype

**DOI:** 10.64898/2026.08.14.26359722

**Authors:** Emma K. Wiener, Rocio Rius, Carlos Dominguez Gonzalez, Arastoo Vossough, Matthew T. Whitehead, Roshini Abraham, Amrita Basu, Nicole Debruyne, Lan Lin, Benjamin L. Prosser, Alex J. Felix, Asako Takanohashi, Kathleen E. Sullivan, Devi Priyanka Maripuri, Amy Pizzino, Kaley Arnold, Alyssa Bryan, Francesco Gavazzi, Mariko Bennett, Sarah E. Hopkins, Brenda Banwell, Vinodh Narayanan, Lindsay Higdon, Kathleen Graveran-Perez, Casey Toback, Michael R. Sperling, Christina A. Gurnett, Noémie Hamilton, Clare E Bryant, Scott W. Canna, Edward M. Behrens, Cas Simons, Adeline Vanderver

## Abstract

**Background:** Monogenic autoinflammatory disorders arise from genetic defects that pathologically activate innate immunity. IRAK4, a serine/threonine kinase in the Myddosome pathway, mediates IL-1 and Toll-like receptor signaling, driving proinflammatory cytokine and type I interferon responses. While biallelic loss-of-function *IRAK4* variants cause an immunodeficiency, recent reports implicate biallelic *IRAK4* variants in severe neuro- and systemic autoinflammation (NASA). We investigated a child with a similar phenotype and screened unsolved autoinflammatory leukoencephalopathies in the Myelin Disorders Biorepository Project (MDBP).

**Methods:** Individuals with unexplained autoinflammatory leukoencephalopathy and no unifying molecular diagnosis were identified in the Myelin Disorders Biorepository Project (MDBP), and genome sequencing was reanalyzed to prioritize rare, protein-altering and splice-affecting variants. Candidate variants and their splicing consequences were interrogated with short-read and targeted long-read RNA sequencing, benchmarked against control PBMC and normal-tissue transcriptomes. Nonsense mediated decay of transcripts was also assessed. Clinical, genetic, and treatment data were extracted by standardized deep phenotyping, and brain MRI was reviewed in consensus by two pediatric neuroradiologists.

**Results:** We identified six patients from five unrelated families with biallelic, rare *IRAK4* variants presenting with severe, persistent autoinflammation without immunodeficiency. Variants included two homozygous and three compound heterozygous changes. All patients had a concordant clinical and radiologic syndrome: episodic, waxing–waning encephalopathy with refractory seizures; neuroimaging showed transient white matter edema that evolved to gliosis, superimposed on marked calcifications and ensuing cerebral atrophy. Biomarkers indicated neuroinflammation and anemia in all cases. Median age at neurologic symptom onset was 12.96 years (IQR 9.44). Immune-suppressive therapies achieved partial benefit, but most patients had ongoing seizures, persistent neuroinflammation, and progressive disease, and without treatment, loss of life.

**Conclusion:** In these six patients, a strongly concordant clinical and radiological phenotype emerges of *IRAK4*- mediated autoinflammation, expanding the phenotypic and mutational spectrum of *IRAK4*-related disease. Further studies are needed to define mechanisms and optimal treatments.

## Introduction

The innate immune system is a critical first line of defense against infection and injury, but when dysregulated can drive severe autoinflammation. Monogenic autoinflammatory disorders are a heterogeneous group of conditions caused by genetic defects that pathologically activate innate immune signaling, often resulting in both systemic and neurologic deficits.

Interleukin-1 receptor–associated kinase 4 (IRAK4) is a serine/threonine kinase that is essential to signaling through the interleukin-1 (IL-1) receptor family and many Toll-like receptors (TLRs)^1,2^. IRAK4 contains two principal functional regions: an N-terminal death domain that serves a scaffolding function, and a C-terminal kinase domain that phosphorylates IRAK1/2 to propagate downstream signaling. When IL-1 receptors (activated by IL-1β and IL-18) and TLRs (activated by pathogen-associated molecular patterns, PAMPs) undergo activation and dimerization, they trigger recruitment of MyD88, IRAK4, and IRAK1/2 to form a supramolecular organizing center known as the myddosome^3–5^ (**Figure 1**). IRAK4 uniquely autophosphorylates and dimerizes, facilitating efficient recruitment and phosphorylation of IRAK1/2, which is key to Myddosome formation. The Myddosome orchestrates the inflammatory response through transcription factors such as nuclear factor κB (NFκB), regulating mRNA stability, and modulating metabolic responses to inflammation^6,7^. These upstream inflammatory events induce the NOD-like receptor protein 3 (NLRP3) inflammasome, which further amplifies inflammation via activation of IL-1β, IL-18 and Gasdermin D which induces pyroptotic lytic cell death. Both IRAK4 scaffolding and kinase activities contribute to stable Myddosome formation^8^. Inhibition of IRAK4 kinase activity reduces—but does not abolish—Myddosome signaling, indicating that the death-domain scaffold function can partially sustain signaling independently of kinase activity.

**Figure 1:**
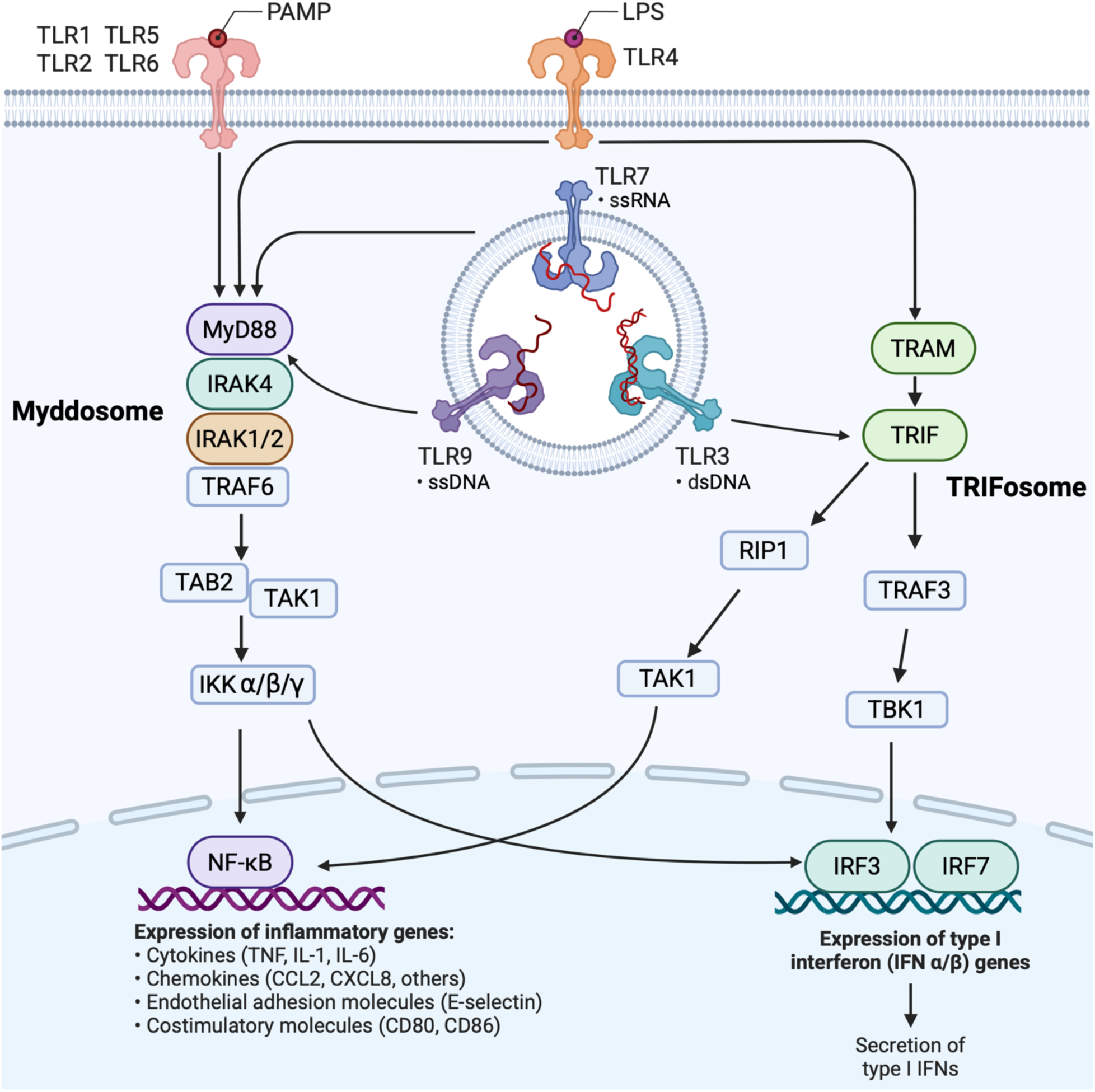
Schematic of the IRAK4 signaling pathways triggered based on different TLR receptors surface or endosomal. The left-hand pathway is the Myddosome pathway that primarily drives inflammatory cytokine production. The right-hand pathway in the non MyD88 pathway also known as the TRIFosome that primarily drives type 1 interferon production.

The “TRIFosome” is a pathway closely related to the Myddosome, triggered by TLR4 and TLR3, that enhances interferon-stimulated genes and increases interferon production^9,10^ (**Figure 1**). Effective IRAK4 signaling also initiates negative-feedback circuits that help restrain the magnitude and duration of proinflammatory responses downstream of TLRs and IL-1Rs^1^. Further, IRAK4 is controlled through alternative splicing, likely playing a role in regulation of autoinflammation. IRAK4 exhibits a variety of alternative splicing patterns in normal tissue which alters the abundance of the resultant functional protein^11^.

Dysregulated or excessive IRAK4 signaling has been implicated in chronic inflammation in diseases such as rheumatoid arthritis^12^, systemic lupus erythematosus^9^, and psoriasis, and it has been linked to oncogenic proliferation in myelodysplastic syndromes and acute myeloid leukemia^13,14^. Consequently, several IRAK4 inhibitors have been developed as potential therapeutics^15^.

Haploinsufficiency of *IRAK4* causes a well-described immunodeficiency^16,17^. Biallelic loss-of-function variants—most commonly nonsense variants that abolish IRAK4 protein—result in an inability to mount effective innate response and predispose affected infants to life-threatening pyogenic infections. A few missense variants have been shown to have allosteric effects that impair Myddosome formation and reduce signaling rather than causing complete loss of protein^18^.

By contrast, recent reports suggest that biallelic hypomorphic *IRAK4* variants can produce a distinct phenotype of severe neuro- and autoinflammation with splenomegaly and transfusion-dependent anemia (NASA syndrome)^19^. Separately, elevated IRAK4 expression and inflammasome-related gene signatures were observed in hippocampal tissue from patients with refractory epilepsy^20^ and overexpression of IRAK4 increased neuronal excitability and seizures in experimental models, whereas IRAK4 inhibition reduced seizures in these epileptic mice^20^. Additionally, a recent publication implicating *IRAK2* mutations in immune dysregulation featured six patients with a homozygous deletion of most of the *IRAK2* death domain with two subjects exhibiting immunodeficiency and four having autoinflammation, similar to the variable phenotypes seen in *IRAK4*-associated disease^21^.

Given these diverse roles of IRAK4 in host defense, inflammation, and neurobiology, we investigated unsolved autoinflammatory leukoencephalopathies in the Myelin Disorders Biorepository Project (MDBP) to identify individuals with similar IRAK4-related disease manifestations. We present here a cohort with biallelic IRAK4 variants who have the autoinflammatory phenotype, but who display an expanded phenotype and genotype spectrum, compared to the original report.

## Methods

### Cohort Recruitment

Probands identified were recruited as affected individuals to the Myelin Disorder Biorepository Project (MDBP) managed by the Leukodystrophy Center of Excellence at The Children’s Hospital of Philadelphia. Clinical records and clinically ordered or research-generated genome and exome sequences were stored for reanalysis in the MDBP’s data system.

### Genome Analysis and Variant Identification

Genome sequencing (GS) data for these cases were processed as part of a MDBP reanalysis initiative in collaboration with the Centre for Population Genomics (CPG) Australia. GS data processing was performed following the DRAGEN GATK best practices pipeline. Reads were aligned to the hg38 reference genome using Dragmap (v1.3.0). Cohort-wide joint calling of single-nucleotide variants (SNVs) and small insertion/deletion (indel) variants was performed using GATK HaplotypeCaller (v4.2.6.1) with “--dragen-mode” enabled^22^. Variants were annotated using VEP 110^23^. Structural variant (SV) calling was performed using GATK-SV^24^. The resulting callset was loaded into CPG’s CaRDinal seqr instance for analysis. Variants were filtered for only rare variants gnomAD v4 AF <0.001 and for exonic, protein-altering variants, and intronic variants with spliceAI scores >0.1. The resulting set of variants were manually reviewed for clinical relevance. Transcript predictions were assessed in Ensembl^25^ (version 116-June 2026) and isoform expression data was interrogated in the GTEX Portal^26^. The data referenced in this manuscript were obtained from the GTEx Portal on 06/30/26 accession number phs000424.v10.p2.

### Clinical Data Extraction

Patients consented to MDBP undergo deep clinical phenotyping. Medical records are used as source documentation. Data on demographics, genotype, age at disease onset, clinical manifestations, and chronological age at the last evaluation are derived from records. Key medical events, including age at presentation and involvement of systemic complications, are extracted using a standardized approach and word bank. For each clinical finding, an age of onset was calculated, or date approximation was used depending on data availability (for example, if only month or season was mentioned, the first day of the regarding period was considered). A symptom or event was considered noted if it was mentioned in their medical records. A symptom or event was considered not noted if a condition was explicitly excluded based on documentation in the medical record. A symptom or event was considered unknown if not mentioned in the medical record. Review of brain magnetic resonance imaging (MRI) was performed by two pediatric neuroradiologists in consensus.

### Short Read RNA Sequencing

Short-read targeted gene specific RNA sequencing was done through LabCorp. Next Generation Sequencing was performed on an Illumina instrument using TruSeq Stranded Total RNA library (Illumina) generated from the submitted whole blood. Sequencing data was aligned with HISAT2 and analyzed using StringTie that allows detection of alterations in transcript ratios and splice site usage^27^. Using the counts associated with each gene the test sample was compared with a set of tissue-specific reference samples to determine a Z-score for assessment of likely significance and the expression change compared to controls.

### Long Read RNA Sequencing

RNA was obtained from 2 patient PBMC samples, 9 additional control PBMC samples, and 7 normal human tissues (blood, corpus callosum, spinal cord, spleen, colon, lung, and stomach) purchased from Clontech (Takara Bio).

Reverse transcription was performed on approximately 500 ng of total RNA using Maxima H minus reverse transcriptase (Thermo Fisher, EP0752), dNTP (Invitrogen, 18427088), template-switching oligo (TSO) (IDT), and RNase inhibitor (Promega, N2615) at 42°C for 90 min, followed by enzymatic inactivation at 85 °C for 5 min. PCR amplification was performed with KAPA HiFi HotStart ReadyMix (Fisher Scientific, 50-196-5217) using the following program: 95°C initial denaturing for 3 min, followed by 13 cycles of 98 °C for 20 s, 67°C for 20 s, 72°C for 5 min, and a final extension at 72°C for 8 min. Amplified cDNA was purified using 0.7X SPRIselect beads (Beckman Coulter, B23318). Quantification was performed using a NanoDrop 2000 Spectrophotometer and Agilent 4200 TapeStation. TEQUILA probes targeting 3190 genes implicated in rare childhood disorders, including IRAK4, were synthesized as previously described ^28,29^. Approximately 500 ng of amplified cDNA was denatured at 95°C for 10 min, then incubated with 100 ng of TEQUILA probes for 12-16 h at 65°C. The cDNA mixture was then incubated with M-270 streptavidin beads (Invitrogen, 65306) for 45 min at 65°C. Heated and room temperature washes were performed as described in the IDT xGen Lockdown protocol. Post-capture on-bead cDNA amplification was performed with KAPA HiFi HotStart ReadyMix using the following program: 95°C initial denaturing for 2 min, followed by 16 cycles of 98 °C for 20 s, 67°C for 20 s, 72°C for 5 min, and a final extension at 72°C for 8 min. Amplified cDNA was purified using 0.7X SPRIselect beads (Beckman Coulter, B23318). Quantification was performed using a NanoDrop 2000 Spectrophotometer and Agilent 4200 TapeStation.

Nanopore libraries were prepared from 200 ng of amplified cDNA following the standard ONT SQK-LSK109 (whole transcriptome, Clontech tissues) or SQK-NBD114 (targeted, PBMCs) protocol. Briefly, the cDNA products underwent end-repair and dA-tailing with the NEBNext Ultra II End Repair/dA-Tailing Module (NEB, E7546) by incubating at 20 °C for 5 min and 65 °C for 5 min, then were purified with 1X AMPure XP beads (Beckman Coulter, A63881). For SQK-NBD114, end-prepped cDNA was incubated with the provided barcodes for 20 min at room temperature and stabilized with EDTA, then purified again. Adapter ligation was carried out at room temperature for 10 min using NEBNext Quick T4 DNA ligase (NEB, E6056). Barcoded samples were pooled, and the resulting library was purified with 0.4X volumes of AMPure XP beads (Beckman Coulter, A63881) and the supplied Ligation Adapter and Short Fragment Buffer, then eluted with the supplied Elution Buffer. 20fmol of the final cDNA library was sequenced on either an R9.4.1 flow cell (FLO-MIN106) or an R10.4.1 flow cell (FLO-PRO114M).

Basecalling of raw nanopore data was performed using super-high accuracy dorado (v.0.9.0). Basecalled reads were aligned to the GRCh38/hg38 primary human reference genome using minimap2 (v2.25) with parameters: ‘-ax splice -ub --secondary=no -k 14 -w 4’. Novel and annotated transcripts were quantified using AMALGAM.

### Nonsense Mediated Decay (NMD) Inhibition Studies

Lymphoblastoid cell lines from Patient 4 and 5 and control cell line GM12878 were treated with 300nM of SMG1 inhibitor (MedChemExpress, HY-124719) or DMSO for 4 hours (n=3 replicates per condition). RT-qPCR was performed using primers targeting IRAK4 constitutive exon 6 (F: 5’-AACACAACTGTGGCAGTGAAG-3’, R: 5’-AAGTTTTCATGTTGACACTTTGCC-3’) and housekeeping gene HPRT1 exon 2 (F: 5’-CCTGGCGTCGTGATTAGTGA-3’, R: 5’-CGAGCAAGACGTTCAGTCCT-3’). For each replicate, ΔCt was calculated as Ct(IRAK4) − Ct(HPRT1), and ΔΔCt as ΔCt(SMG1i) − median ΔCt(DMSO) for that cell line; fold change was calculated as 2^−ΔΔCt. Statistical comparisons of ΔΔCt between cell lines were performed using a one-way ANOVA followed by Dunnett’s post hoc test, with GM12878 as the control group and significance set at α=0.05.

### In silico Variant Modelling

Structural modelling was performed using UCSF ChimeraX^30^ and the MyD88-IRAK4-IRAK2 death-domain complex structure PDB 3MOP^31^. MyD88 chains A-F, IRAK4 chains G-J, and IRAK2 chains K-N were retained. IRAK4 variants p.Asp27Ala and p.Ala97Val were introduced into IRAK4 chains G-J by side-chain substitution using the backbone-dependent rotamer library^31,32^ with backbone coordinates kept fixed. Local contacts, steric clashes, hydrogen bonds, and inter-chain distances around the variant residues were assessed in ChimeraX and compared with the wild-type model. DynaMut2^33^ was used as an independent estimate of local stability change using representative IRAK4 chain I. Predicted ΔΔG values were recorded for p.Asp27Ala and p.Ala97Val. Conservation was assessed using a multiple sequence alignment of vertebrate IRAK4 orthologues from UniProtKB.

## Results

### Cohort description

Six individuals from five unrelated families were identified with biallelic variants in *IRAK4*, four females and two males. Four individuals had compound heterozygous variants (Patient 1, Patient 6 and Patients 2 & 3 who were siblings) and two individuals had homozygous variants (Patient 4 and Patient 5).

### Neurological disease description

Patients presented with both systemic and neurological inflammatory features summarized in **Table 1**. In three cases, the first presenting feature was systemic, whereas in the other three, their first presenting symptom was neurological. In all patients the first neurological symptom was a seizure with a median age of 12.96 (IQR 9.44). The systemic presenting features differed between cases, with a median age of systemic disease onset of 9.96 (IQR 13.7).

**Table 1:** *IRAK4 (NM_001114182.3)* Variants and Clinical Manifestations of disease.

|  | Patient 1 | Patient 2 | Patient 3 | Patient 4 | Patient 5 | Patient 6 |
| --- | --- | --- | --- | --- | --- | --- |
| **IRAK4 allele 1 HGVS c.** | c.539_540del | c.877C>T | c.877C>T | c.161+892A>G | c.333del | c.877C>T |
| **IRAK4 allele 1 HGVS p.** | p.Phe180Ter | p.Gln293Ter | p.Gln293Ter | p.? | p.Leu112TyrfsTer7 | p.Gln293Ter |
| **IRAK4 allele 2 HGVS c.** | c.80A>C | c.162-217C>T | c.162-217C>T | c.161+892A>G | c.333del | c.290C>T |
| **IRAK4 allele 2 HGVS p.** | p.Asp27Ala | p.? | p.? | p.? | p.Leu112TyrfsTer7 | p.Ala97Val |
| **Age at neurological symptom onset (years)** | 11-15 | 16-20 | 11-15 | 6-10 | 21-30 | 0-5 |
| **Age at systemic symptom onset (years)** | 16-20 | 0-5 | 11-15 | 0-5 | 21-30 | 0-5 |
| **MRI abnormalities** |  |  |  |  |  |  |
| Abnormal Calcifications | ++ | ++ | ++ | ++ | ++ | + |
| Transient or migrating white matter signal abnormality and edema | + | + | + | + | + | + |
| White matter signal abnormalities | Multifocal | Multifocal | Multifocal | Multifocal | Multifocal | Multifocal |
| Areas of contrast enhancement | + | — | + | + | + | + |
| **Inflammatory Markers** |  |  |  |  |  |  |
| Elevated Sedimentation Rate | + | + | + | + | + | + |
| Serum Cytokines | Not done | Not done | Not done | Elevated IFN-gamma, IL-6, IL-8, IL-10, TNF-a | Elevated IL8 | Elevated IFN-gamma, IL-6 |
| CSF neopterin | 282 | Not done | Not done | Persistently >200 | Normal | 115 |
| CSF Protein | High | High | High | High | High | High |
| CSF Cytokines | Not done | Not done | Not done | IL-6, IL8, Interferon gamma | Elevated IL-6 | Elevated IL-6 |
| **Neurologic features** |  |  |  |  |  |  |
| Refractory Seizures | + | + | + | + | + | + |
| Visual loss | + | + | + | + | + |  |
| Cognitive decline | + | + | + | + | + | + |
| Behavioral/psychiatric changes | + | + | + | + | + | + |
| Headaches | + | + | + | + | + | + |
| Motor symptoms | - | - | - | + | + | - |
| **Systemic Features** |  |  |  |  |  |  |
| Anemia | + | + | + | + severe | + | + |
| Hepato/Splenomegaly | - | Mild Hepato-/Splenomegaly | Mild Hepato-/Splenomegaly | Palpable Hepato-/Splenomegaly | Mild Hepatomegaly | Mild Hepatomegaly |
| Gastrointestinal Inflammation | + | + | + | + | - | - |
| Thrombocytopenia | + | + | + | - | - | + |

+ present ++ severe

Four out of the six patients had a clinically focal to bilateral tonic-clonic seizures for their first episode but then went on to have predominantly focal impaired consciousness seizures that often remained in status despite multiple agents. Some patients experienced subclinical seizures detected by continuous EEG. Episodes were most often accompanied by a prolonged altered mental state, transient visual loss 5/6 and extreme psychomotor agitation. Five patients also complained of severe headaches both during episodes and outside of them. All cases showed progressive cognitive decline as refractory seizure episodes worsened. In those who experienced visual changes the changes progressed from transient to permanent visual loss in 4/6 cases. All cases demonstrated psychiatric symptoms and behavioral changes after disease onset; 4/6 developed aggressive behavior and outbursts, 4/6 extreme anxiety, 3/5 depressed mood, 1/6 obsessive behavior, 1/6 hyperactivity and inattention, 2/6 hallucinations (visual and auditory). There was little motor involvement early in the disease course aside from hyperreflexia present in 5/6 cases and two cases had hypertonia with altered gait.

### Systemic disease description

Systemic manifestations of disease varied between individuals. Four of the six patients had inflammatory gastrointestinal symptoms and two of the four received diagnoses of atypical inflammatory bowel disease (IBD). One patient had hypothyroidism and type 2 diabetes (not confirmed to be autoimmune); one patient had occasional skin rashes. All patients had unexplained chronic microcytic anemia of varying severity; in one case this was transfusion-dependent anemia. Four out of six cases had thrombocytopenia. One patient had rheumatoid factor negative polyarticular juvenile idiopathic arthritis and a mild Von Willebrand-like bleeding disorder. Three of the six patients had hepato-/splenomegaly noted, and two patients had only mild hepatomegaly accompanied by elevated liver enzymes.

### Inflammatory markers and Investigations

Inflammatory investigations were consistent with neurological and systemic inflammation (**Table 1**). Erythrocyte sedimentation rate (ESR) was elevated persistently in 5/6 patients but only transiently in Patient 6. Patients 4, 5 and 6 had serum cytokine panels and interferon activity testing; Patient 4 and Patient 6 had elevated interleukin (IL)-6 and interferon gamma, with Patient 4 also having elevated IL-8, IL-10 and TNF-alpha. In Patient 5 only IL-8 was elevated. Patient 4 had an elevated monocyte type 1 interferon signature but Patients 5 and 6 did not.

Cerebrospinal Fluid (CSF) testing was done in all patients and all showed persistently elevated protein, and 4/6 patients had elevated lymphocytes. 3/6 patients (Patients 1, 4 and 6) had elevated neopterin, although this was not tested in Patients 2 and 3.

Patients 4, 5 and 6 had extensive cytokine testing on CSF which was not performed on the other patients. All three patients had elevated CSF IL-6. Patient 6 had one CSF test with elevated IL-8 at first presentation and Patient 4 additionally had persistently elevated IL-8, IL-10, TNF-alpha, and IFN gamma. Patients 2, 5 and 6 had brain biopsies that all showed reactive gliosis with hypercellularity/lymphocytic infiltrate. In Patient 2 mild spongiform changes were noted and in Patient 5 mild capillary hyperplasia.

### Neuroimaging features

Neuroimaging is shown in **Figure 2A and B** and findings are detailed for each patient in **Table 1**. In each patient serial neuroimaging displayed an evolving course, this progression is shown in series of images for Patient 5 in **Figure 2B**. Calcifications visible on CT variably involved the basal ganglia, subcortical white matter, and periventricular white matter. Axial FLAIR images and postcontrast T1-weighted images show waxing and waning migrating white matter signal abnormality, edema, and gyriform/leptomeningeal contrast enhancement, at times with some mild residual gliosis and mild focal volume loss

**Figure 2:**
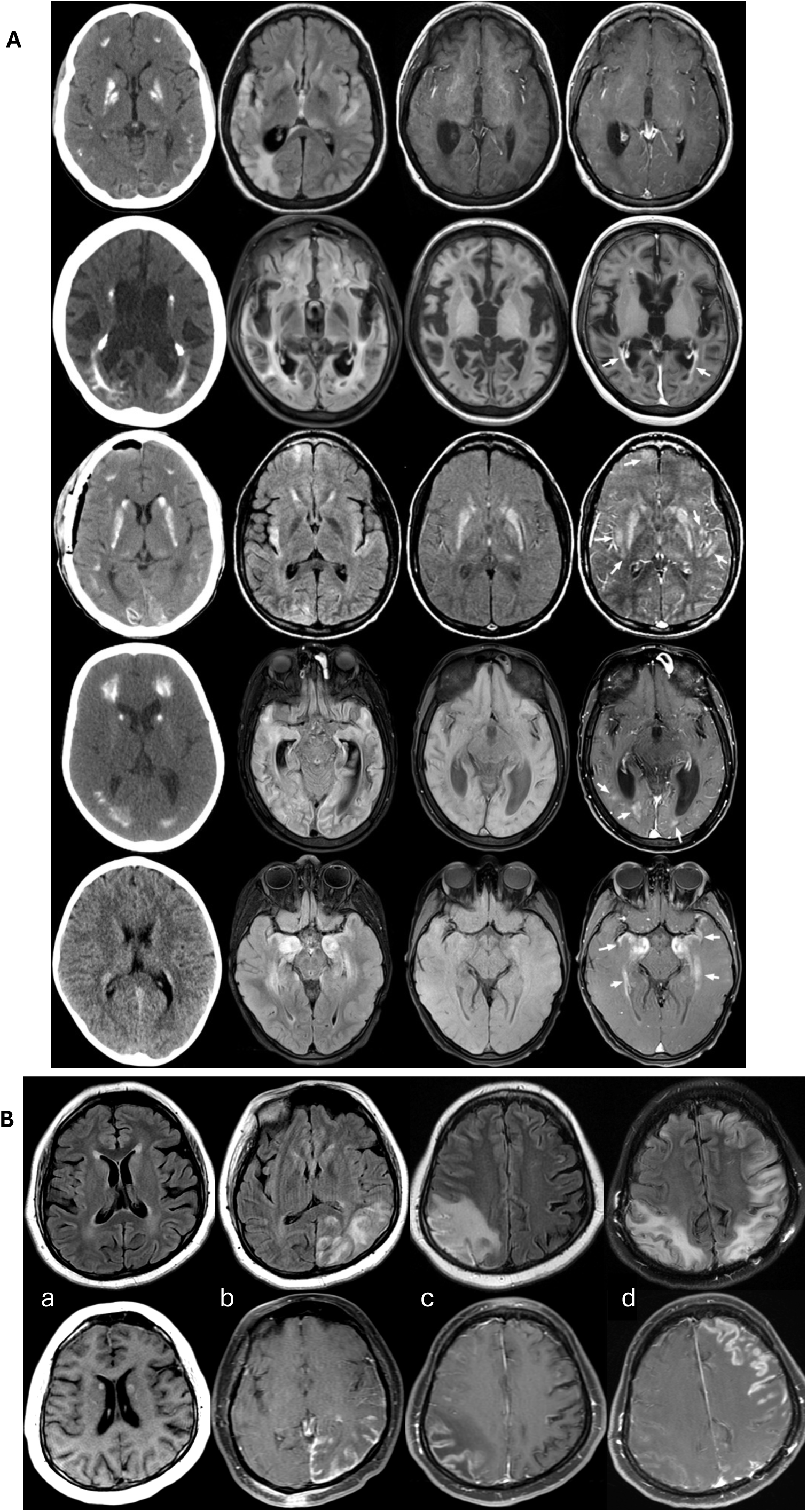
Neuroimaging findings in patients with biallelic IRAK4 variants and autoinflammation. **(A)** Representative imaging of salient features in IRAK4 cases. Row 1 -Patient 3, Row 2 – Patient 1, Row 3 – Patient 2, Row 4 – Patient 4, Row 5 – Patient 6. The first column shows CT images showing abnormal calcifications in the brain in all patients, variably involving the basal ganglia, subcortical white matter, and periventricular white matter. Axial FLAIR imaging shows multifocal signal abnormalities (column 2) in the brain involving both gray and white matter. Axial precontrast T1 (column 3) and axial postcontrast T1 (column 4) show areas of contrast enhancement (arrows), except in row 1 that did not have enhancement at the time of the MRI. **(B)** This shows the progression of imaging findings in one patient (Patient 5) at four timepoints **(a)** at disease onset, **(b)** one year later, **(c)** 2 years later and **(d)** 6 years later. The top row shows axial FLAIR images and bottom row shows postcontrast T1-weighted images. The images show waxing and waning migrating white matter signal abnormality, edema, and gyriform/leptomeningeal contrast enhancement, at times with some mild residual mild gliosis and mild focal volume loss.

### Disease Course and Treatment

All patients showed significant progression of disease severity after onset of seizures. Multiple immune modulatory agents were used in all cases. Patients 1, 2, and 3 were seen before targeted immunomodulatory therapies were widely available; treatment was therefore limited to methotrexate for arthritis, IVIG, and steroids. All three of these individuals had continuous disease progression with some transient improvement from steroids but continued to decline and passed away 0.5-8 years after seizure onset.

In Patients 4, 5 and 6 multiple agents were required to address their systemic manifestations (anemia, endocrine and GI inflammation) and neurological manifestations (seizures, developmental regression, cognitive decline, vision loss, psychiatric and motor symptoms). In all three patients, multiple immune modulatory agents were required, in addition to agents such as antiepileptics, antipsychotics and other symptom specific agents. Seizure control was a significant part of disease control and seizures became refractory without the addition of immune modulatory agents. Inflammatory markers, MRI changes in addition to symptom control were used to monitor treatment success and direct therapy changes and additions. In all three patients, steroids were effective in minimizing symptoms; however, due to the high doses required and concerns about the side effects of chronic use, steroids were weaned as far as possible in favor of targeted agents. Based on the inflammatory markers in CSF and serum (cytokine and interferon testing) agents such as siltuximab, tociluzumab, baricitinib and anifrolumab were initiated. These agents showed some success in improving symptoms and reducing measurable inflammation: CSF protein, Neopterin, CSF and serum cytokine and interferon testing. Additionally, cyclophosphamide was used in two patients, and in one of the two it was stopped in favor a transition to mycophenolate mofetil.

### Genetic Findings

All six individuals were found to have biallelic variants in *IRAK4* (**Table 1 and Figure 3**) with confirmed biparental inheritance. Two individuals had homozygous changes: Patient 5 with c.333del (p.Leu112TyrfsTer7); and Patient 4 with c.161+892A>G a deep intronic variant with SpliceAI score of 0.85 predicted to produce several alternative transcripts. This splicing effect was confirmed on clinical short-read RNA-seq indicating a complete absence of the canonical exon 2-3 junction in the main *IRAK4* transcript (NM_016123.4). This results in an abnormal transcript containing 2055 bases of intron 2 that leads to a premature termination codon (p.ArgThrfs*18). However, 30% of all *IRAK4* transcripts represented alternative isoforms (NM_001145257.2, NM_001145256.2 and NM_001351344.2) which utilize an alternative start codon at amino acid 124 of the full length IRAK4 protein.

**Figure 3:**
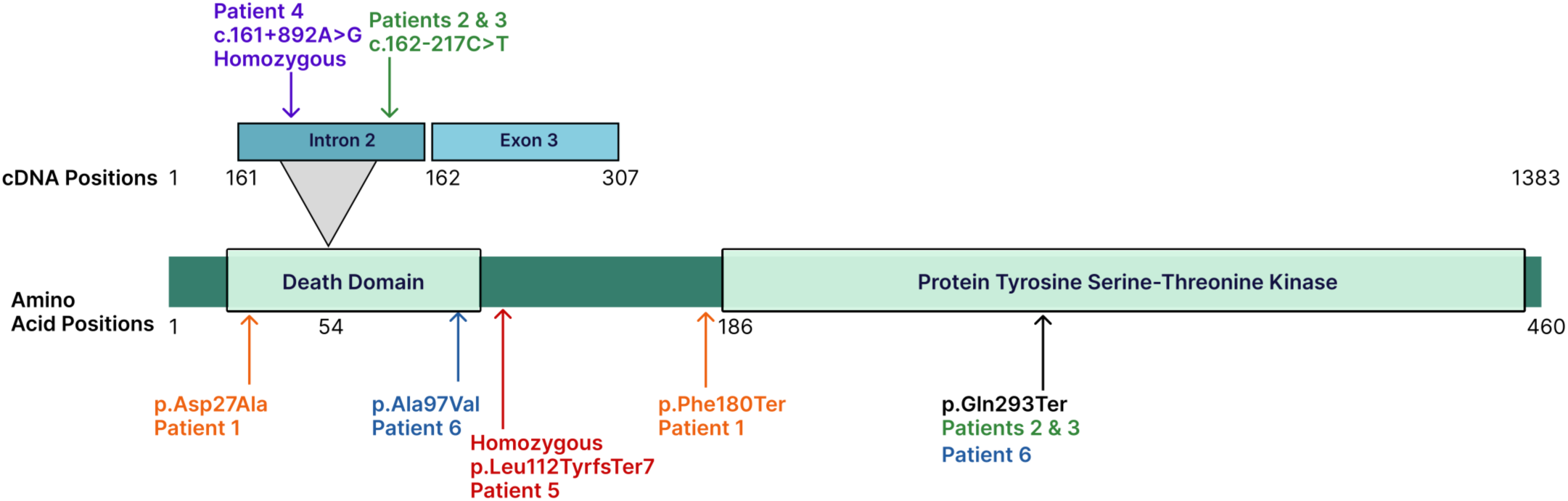
Schematic of IRAK4 indicating the position of each patient’s variant/s. Variants are represented using HGVS p. nomenclature except for the splice variants which are represented with HGVS c. nomenclature. Variants are heterozygous except when listed as homozygous. Variants in Patient 1 are colored orange, Patients 2 & 3 in green, Patient 5 in red, Patient 4 in purple and Patient 6 in blue.

Patient 1 and Patient 6 both had compound heterozygous variants, one null variant and one missense variant in the epitope within the death domain that binds to IRAK1 or IRAK2. Patient 1 had compound heterozygous variants, c.539_540del (p.Phe180Ter) and c.80A>C(p.Asp27Ala), and Patient 6 had c.877C>T (p.Gln293Ter) and c.290C>T (p.Ala97Val). The missense variant p.Asp97Ala additionally has a SpliceAI score for Donor loss of 0.2 and Donor gain of 0.86. These likely cause partial deletion or skipping of exon 3 respectively and therefore might similarly result in utilization of the alternative start codon at amino acid 124.

The last two patients, who were siblings, had one null change: c.877C>T (p.Gln293Ter) and one deep intronic variant c.162-217C>T predicted to produce two cryptic splice sites with splice AI scores of 0.65 and 0.61, respectively. The variant would result in increased usage of this cryptic splice site and resultant increase in the proportion of alternative transcript isoforms ENST00000550386.5 and ENST00000696796.1. In Ensembl^25^ both these transcripts are predicted to undergo NMD, so this variant would result in decreased expression of the canonical functional IRAK4.

Of note c.877C>T (p.Gln293Ter) appears to be a recurrent variant; has an allele frequency of 0.08% in gnomADv4. It is reported in two families in this cohort and multiple published individuals with IRAK4 immunodeficiency^34^.

### Long read RNA sequencing in Patients 4 and 5

To determine the effect of the variants in Patients 4 and 5, on splicing of *IRAK4*, targeted long read nanopore RNA sequencing of 3190 genes implicated in rare childhood disorders, including *IRAK4* was performed on PBMC samples. Given that *IRAK4* is known to have several alternative splicing patterns (**Figure 4 Panel A)**, two control data sets were used for comparison to Patients 4 and 5 (**Figure 4 Panels B & C)**. One set of 9 PBMC control samples had the same targeted long-read nanopore RNA-sequencing. A second control set of previously generated whole transcriptome long read nanopore sequencing data set from 7 normal human tissues (Clontech) (**Figure 4 Panels B & C)**. The latter includes 300 reads overlapping the canonical transcript of *IRAK4*, which is of comparable depth to the targeted PBMC samples in the former control data set.

**Figure 4:**
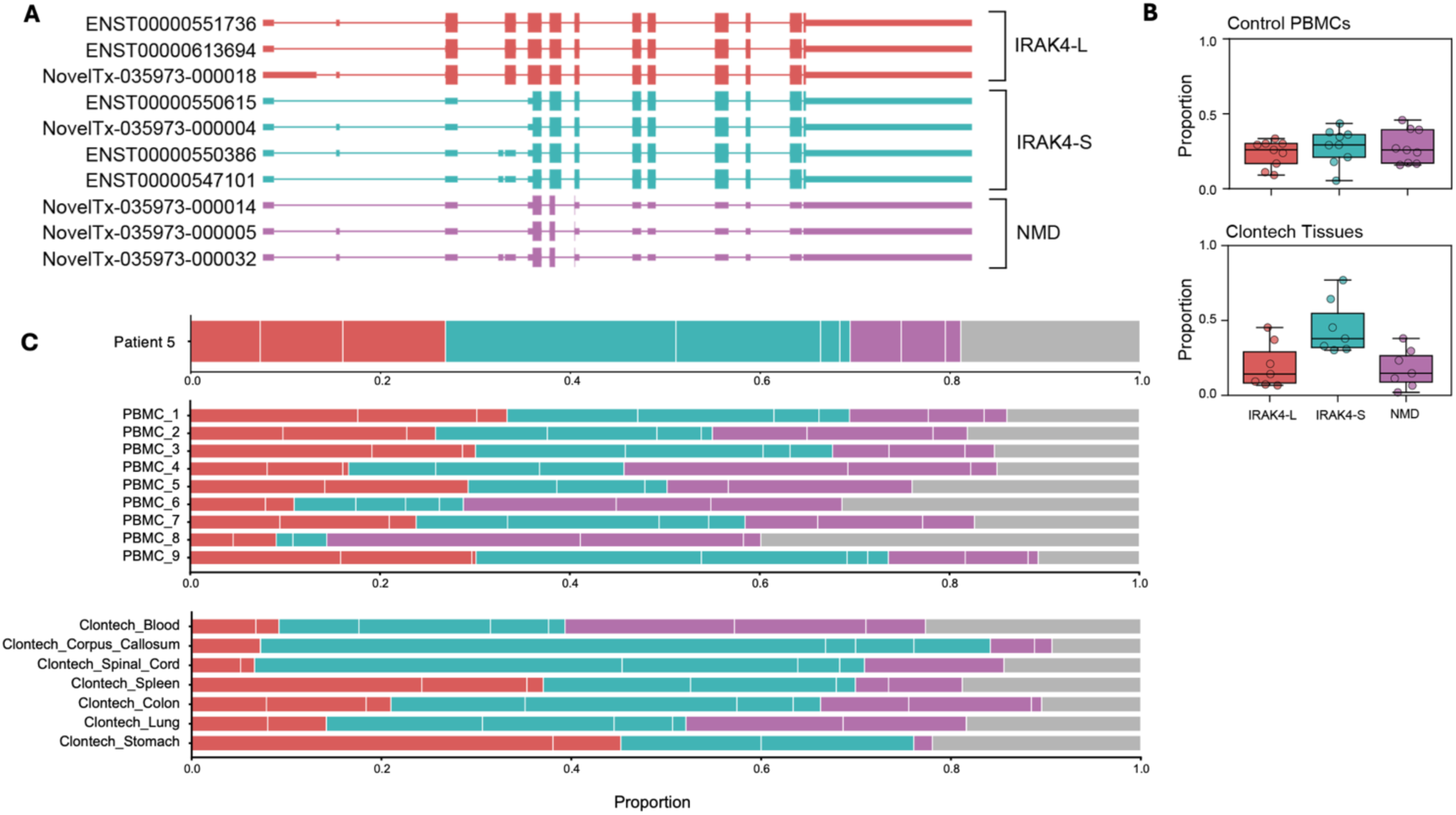
Long Read RNA Sequencing of IRAK4 transcripts in control datasets and Patient 5. **(A)** Structures of the top 10 expressed mRNA isoforms in control PBMCs and Clontech tissues, grouped by predicted coding sequence (red: IRAK4-L, teal: IRAK4-S, purple: NMD). **(B)** Aggregated predicted coding sequence usage in the 9 control PBMCs and 7 Clontech tissues. (red: IRAK4-L, teal: IRAK4-S, purple: NMD). **(C)** Breakdown of predicted coding sequence usage in the 9 control PBMCs and 7 Clontech tissues. (red: IRAK4-L, teal: IRAK4-S, purple: NMD). Marked variability is seen in the proportion of predicted coding sequence usage between individuals and between different tissues.

The variants identified in each patient are shown in **Figure 5 Panels A & C**. Patient 5 possesses a 1 bp deletion in exon 4 which causes no change in splicing. Patient 4 possesses an intronic SNV which creates a cryptic splice acceptor. Both variants are predicted to result in a premature termination codon on transcript isoforms which use the canonical start codon (IRAK4-L), which likely undergo NMD and therefore these patients are unlikely to produce the full length IRAK4 protein.

**Figure 5:**
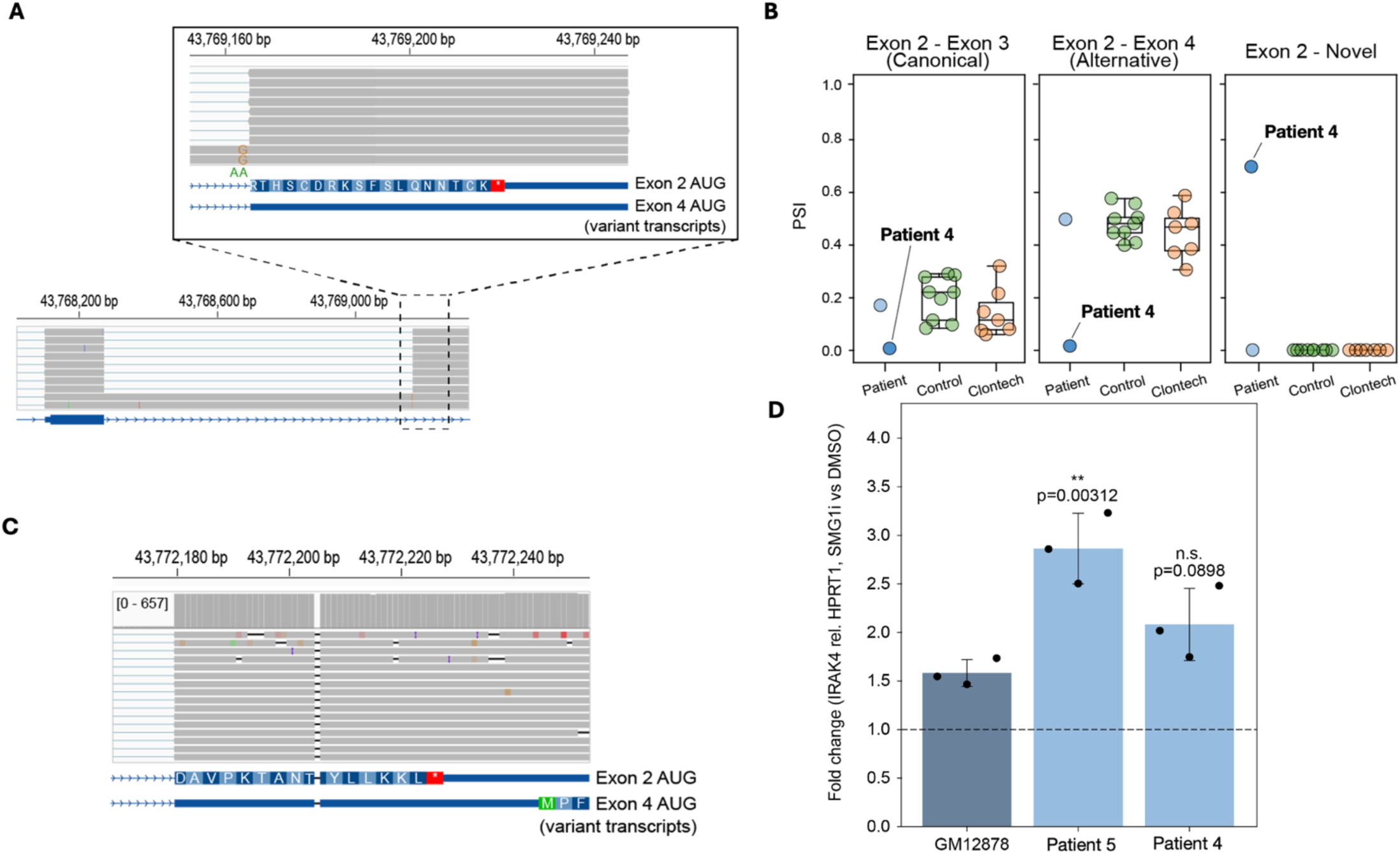
Long Read RNA Sequencing in Patients 4 & 5 and controls. **(A)** IGV screenshot showing the intronic single nucleotide variant in Patient 4, which creates a cryptic splice acceptor in intron 2 and results in a premature termination codon 18 codons downstream. Resulting transcripts are predicted to be degraded by NMD, assuming they use the canonical start codon corresponding to IRAK4-L. Alternatively, if these transcripts utilize the distal start codon corresponding to IRAK4-S, all splicing changes occur within the 5’ UTR. **(B)** Percent spliced-in (PSI) values from patient (blue), control PBMCs green and clontech (orange). PBMCs for the most common annotated junction that uses the intron 2 splice donor (which skips exon 3) as well as the novel junction created by the SNV in Patient 4. **(C)** IGV screenshot showing the 1 bp deletion in exon 4 of Patient 5, which occurs just upstream from the IRAK4-S start codon. This deletion is predicted to result in a frameshift and PTC on transcripts encoding IRAK4-L which are therefore predicted to be degraded by NMD. This deletion is present in the 5’ UTR of transcripts encoding IRAK4-S. **(D)** Fold change of IRAK4 in cells treated with SMG1i versus DMSO. Error bars represent mean fold change ± SD (n = 3 replicates per condition). ** p < 0.01, Patient 5 vs. GM12878 (Dunnett’s test). Significantly more transcripts in Patient 5 undergo NMD compared to controls, Patient 4 has more transcripts undergoing NMD but fewer than Patient 5. In both patients some, but not all, transcripts undergo NMD.

However, both variants occur upstream from the distal start codon (IRAK4-S) and therefore cause changes to the 5’ UTR on the alternatively spliced transcripts. As a result, variant transcripts may either: 1) utilize the canonical start codon, contain a PTC, and therefore are targeted for degradation by NMD, or 2) begin or reinitiate translation at the distal start codon in exon 4 and therefore produce IRAK4-S protein product (**Figure 5 Panel A & C)**.

To assess the level of nonsense-mediated decay of IRAK4 transcripts, we treated patient and control (GM12878) lymphoblastoid cell lines with an SMG1 kinase inhibitor (SMG1i) to inhibit NMD and quantified total IRAK4 mRNA expression with RT-qPCR relative to housekeeping gene HPRT1. In GM12878 cells, IRAK4 expression increases 1.6-fold following inhibition of NMD (**Figure 5 Panel D),** indicating that some degree of NMD occurs at baseline in normal cells. This is consistent with the presence of NMD-targeted transcripts in control PBMCs and Clontech tissues (**Figure 4B**). Patient 5 cells exhibit a significant 2.8-fold increase (p=0.00312, Dunnett’s test vs. GM12878), while Patient 4 cells show a trend toward increased expression (2.1-fold, p=0.0898) that did not reach statistical significance (**Figure 5 Panel D).** These results indicate that a larger proportion of patient *IRAK4* transcripts are degraded by NMD than in control cell lines, and suggest that some, but not all, variant transcripts are targeted by NMD in these patients.

### In silico protein modelling

Protein modeling for the missense variants in Patients 1 and 6 is shown in **Figure 6**. In the 3MOP MyD88-IRAK4-IRAK2 death-domain complex, p.Asp27Ala and p.Ala97Val were both located within the modeled IRAK4 death-domain construct and were not directly embedded in tight inter-chain contacts (**Figure 6 Panel B)**. p.Asp27Ala lies near the N-terminal D27/Q29 loop region (and was associated with loss of local polar/H-bond interactions, consistent with altered loop stabilization (**Figure 6 Panels E & F)**. DynaMut2 predicted a destabilizing effect for p.Asp27Ala (ΔΔG -0.792 kcal/mol). p.Ala97Val showed preserved H-bonding but increased local hydrophobic packing and steric clashes with neighboring IRAK4 residues (**Figure 6 Panels C & D)**. DynaMut2 predicted a smaller destabilizing effect for p.Ala97Val (ΔΔG -0.207 kcal/mol). Overall, these findings suggest that p.Asp27Ala has the stronger predicted structural impact, whereas p.Ala97Val may act through a more localized intra-IRAK4 packing effect.

**Figure 6:**
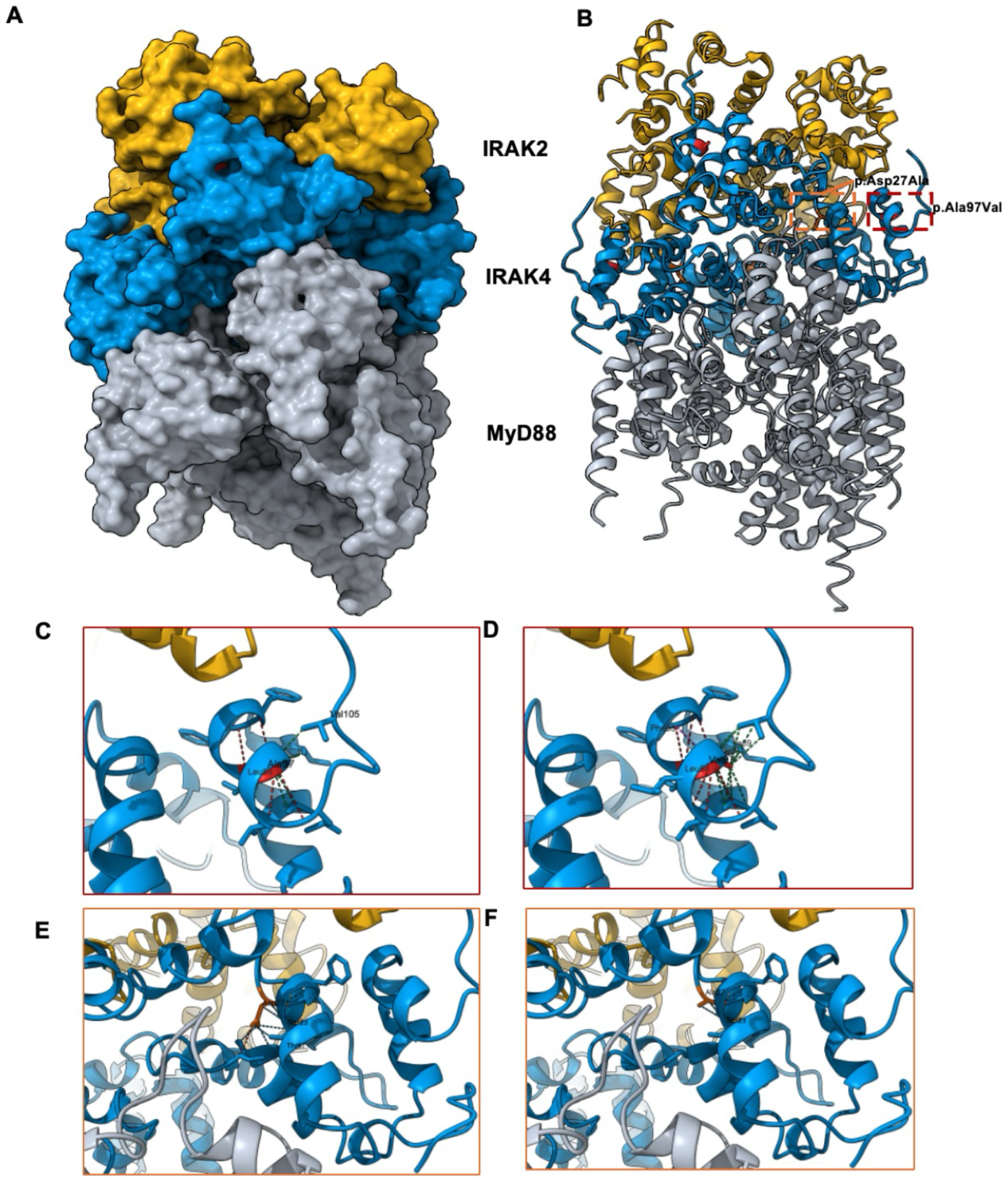
Structural modeling of IRAK4 missense variants in the 3MOP Myddosome complex. **(A)** Surface representation of the assembled Myddosome (PDB 3MOP), colored by protein identity: MyD88 (gray), IRAK4 (blue), and IRAK2 (gold). **(B)** Cartoon representation of the 3MOP complex with the two individual-specific IRAK4 missense sites highlighted on all four IRAK4 protomers (chains G-J): the p.Asp27Ala site in orange and the p.Ala97Val site in red. The boxes mark the regions enlarged in panels C-F and are color-matched to those panel borders. The two variants were identified in different patients and are not modeled as combined. **(C–F)** Close-up views of the local environment at each variant site, comparing wild type with mutant. Predicted interactions are drawn as dashed pseudobonds: hydrogen bonds (red), polar contacts (cyan), hydrophobic contacts (green), and steric clashes (magenta). The substituted side chain is highlighted in red at the p.Ala97Val site and in orange at the pAsp27Ala27 site **(C)** Wild-type Ala97, packing against neighboring hydrophobic residues of the IRAK4 core (e.g. Val105). **(D)** The p.Ala97Val substitution introduces additional hydrophobic contacts and short-range steric clashes within the same core. **(E)** Wild-type Asp27, participating in a local polar/hydrogen-bond network (e.g. Ser23, Thr67). **(F)** The Asp27Ala substitution abolishes the aspartate side-chain contacts, leaving only a local backbone hydrogen bond.

Both affected amino acids were highly conserved across the IRAK4 orthologue alignment AlphaMissense in silico pathogenicity scores were 0.9657 for p.Asp27Ala and 0.5696 for p.Ala97Val (developer-recommended thresholds: ambiguous 0.34–0.564, likely pathogenic > 0.564)^35^. As mentioned above p.Ala97Val also has a high SpliceAI score 0.86. Given the moderate AlphaMissense score and the high predicted splice score this variant may be acting through a splice mechanism and not due to altered amino acid properties.

## Discussion

We present a cohort of 6 individuals from 5 unrelated families with a strongly concordant clinical and radiological phenotype of IRAK4-mediated autoinflammation that expands the phenotypic and mutational spectrum of IRAK4-related disease from previously published cases. The first report of *IRAK4* related NASA syndrome by Cooray et al. described individuals with age of onset from birth – 24months (median 8 weeks and mean 44.5 weeks). This cohort has a median age of neurological onset of 12.96 (IQR 9.44), and median systemic symptom onset of 9.96 (IQR 13.7). This age of onset is markedly higher than the previously published cohort. This later age of onset, especially in the adult patient, poses the question of whether there is a triggering event that causes the onset of the neurological disease in these patients. As this cohort includes individuals treated before targeted immune suppressive agents it also gives a picture of the untreated natural history of this disorder which is severe and fatal in these three historical cases.

The neurological disease observed in the 6 patients was strongly concordant; seizure types were predominantly focal impaired consciousness seizures with associated prolonged altered mental state, agitation, transient visual loss and headaches. Seizure episodes also often remained in status epilepticus and responded very poorly to antiepileptics. In the individuals who did not receive targeted immunomodulation, increasing frequency and severity of these episodes was associated with progressive cognitive and functional decline. Another feature not previously described is the neuropsychiatric mood and behavioral changes after disease onset observed in a number of this cohort. Behavioral and psychiatric manifestations have been reported in later-onset and adult cases of Aicardi-Goutières syndrome^36,37^ and additionally are a reported common side effect of interferon therapy when used for melanoma and Hepatitis C^38^. It is possible that the psychiatric manifestations reported are more prominent in these cases due to the later age of onset of these patients. We did not observe an early motor component in these cases unlike other interferonopathies like where motor disease is prominent^39^, although over time patients uniformly had profound decline in functional status.

In the original cohort, systemic manifestations included recurrent fevers, anemia, hepatosplenomegaly, and transaminitis. These features were also present in this cohort, though fevers and transaminitis appeared less frequently. Most participants additionally experienced gastrointestinal inflammation and thrombocytopenia. Other systemic inflammatory conditions observed included juvenile arthritis, bleeding disorders, hypothyroidism, and diabetes.

Biochemical markers of inflammation were in keeping with what has been previously reported with all cases having elevated ESR and protein in CSF and all cases tested having elevated IL-6 in CSF. Unfortunately, half this cohort were historical, deceased cases who could not be tested for cytokines or interferon. Only one of the three cases tested had persistent monocyte interferon signature, although interferon gamma was also elevated in the serum of Patient 6.

Neuroimaging findings in this cohort were consistent with those described by previous authors^19,40^, although we observed patients with slightly more severe neuroimaging findings than those described previously. Immunosuppressive approaches, including long-term steroids, biologics and alkylating agents, appear to mitigate symptoms and disease progression, relative to untreated disease, which was otherwise progressive and lethal. However, despite significant immunosuppression, some level of inflammatory activity in the intrathecal space appeared persistent, consistent with prior reports^40^. Further work is needed to find a better treatment strategy for this disease.

The variants identified in this cohort expand the known variant spectrum. The previous report of IRAK4-related NASA syndrome^19^ reported compound heterozygous variants with a single null variant and a missense variant. Each has a null variant predicted to undergo NMD and a second missense change. One stabilizes the kinase domain, and one alters the death domain interactions and was thought to result in aberrant Myddosome assembly and disassembly^19^. Our cohort includes two cases (Patients 1 and 6) with similar genotypes as this prior report. *In silico* modeling of the p.Asp27Ala missense variant in Patient 1 causes a similar structural alternation and thus likely similar mechanism to p.Gln29Pro described by Cooray et al. where the variant altered Myddosome assembly and possible dysregulated downstream signaling. The variant in Patient 6, p.Ala97Val, is a missense change that causes steric clashes in the final helix of death domain and is predicted to affect inter-IRAK4 packing which might be altering Myddosome assembly. It might additionally introduce a cryptic splice site resulting in transcripts that are translated using the alternative start codon at amino acid 124 similar to other cases in this cohort with altered splicing patterns. The result of this predicted splicing in being investigated for this patient.

Additionally, in this cohort we also describe different types of genotypes likely resulting in an excessive production of alternative transcripts. We report two cases with homozygous genotypes. Both homozygous variants in Patient 4 and 5 produce several transcripts with termination codons, which were shown to undergo NMD. However these variants are predicted to produce alternative transcripts with a downstream start codon after exons 2/3 that produce a shorter IRAK4 isoform that only includes the kinase domain. Long read RNA sequencing results and the NMD studies support that these homozygous variants result in the preservation of the alternative transcripts predicted to utilize the alternative start codon. In addition to these homozygous variants with loss of the death domain and retention of an alternative kinase only domain, we also report two siblings (Patients 2 and 3) who are compound heterozygous for a single recurrent null and a deep intronic splice variant. Unfortunately, these cases are deceased, and no further investigation is possible. However, from in silico predictors, it seems likely that some transcripts produced would be normally spliced and an increased percentage would undergo NMD resulting in a small amount of functional IRAK4 protein. Similarly to the other cases, enough IRAK4 protein is produced with signaling capability to prevent the infantile immunodeficiency phenotype, but innate immune control is lost, and autoinflammation ensues.

Little is known about the role of alternative isoforms in IRAK4. These alternative transcripts are produced in normal tissue as well, but the proportion of them that are translated under normal conditions, and the role of this smaller protein physiologically, is not known. The abundance of these alternative transcripts clearly differs between tissues and cell types as seen both in the long read RNAseq control datasets as well as GTEX data^26^, and between individuals. Research into MDS and AML^13,14^ indicates that an alteration of this balance can drive oncologic progression and possibly, a different imbalance that is seen in these patients, may be driving dysregulation of innate immunity.

Given that some of these cases have a later onset than previously published cases, there is a question of whether there was a trigger that altered protein expression, isoform usage or signaling pathway balance towards dysregulation, resulting in the disease onset. Patient 6 who has the latest disease onset following normal disease-free development prior, had a mild traumatic brain injury preceding neurological symptom onset. The other cases do not demonstrate an identifiable trigger to progressive brain disease. Further work will be needed to understand the factors and forces that control IRAK4 mediated immune function.

This report shows both an expansion of the described clinical disease as well as novel genotypes for IRAK4 related NASA syndrome. We describe four genotypes that appear to be causing disease not by protein alteration from missense variants but by an imbalance of alternatively spliced *IRAK4* transcripts. While the immunological mechanism of disease remains uncertain in IRAK4 related NASA syndrome, this cohort expands our understanding of the scope of IRAK4 related neuro-and auto-inflammation.

## Ethics Declaration

Ethics approval was provided by Children’s Hospital of Philadelphia IRB #14–011236. All participants signed informed consent to the Myelin Disorders Biorepository Project and for publication.

## Data Availability

All data produced in the present study are available upon reasonable request to the authors.

